# Evidence for gastrointestinal infection of SARS-CoV-2

**DOI:** 10.1101/2020.02.17.20023721

**Authors:** Fei Xiao, Meiwen Tang, Xiaobin Zheng, Chunna Li, Jianzhong He, Zhongsi Hong, Siwen Huang, Zhenyi Zhang, Xianqi Lin, Zhaoxiong Fang, Renxu Lai, Shoudeng Chen, Jing Liu, Jin Huang, Jinyu Xia, Zhonghe Li, Guanmin Jiang, Ye Liu, Xiaofeng Li, Hong Shan

## Abstract

The new coronavirus (SARS-CoV-2) outbreak originating from Wuhan, China, poses a threat to global health. While it’s evident that the virus invades respiratory tract and transmits from human to human through airway, other viral tropisms and transmission routes remain unknown. We tested viral RNA in stool from 73 SARS-CoV-2-infected hospitalized patients using rRT-PCR. 53.42% of the patients tested positive in stool. 23.29% of the patients remained positive in feces even after the viral RNA decreased to undetectable level in respiratory tract. The viral RNA was also detected in gastrointestinal tissues. Furthermore, gastric, duodenal and rectal epithelia showed positive immunofluorescent staining of viral host receptor ACE2 and viral nucleocapsid protein in a case of SARS-CoV-2 infection. Our results provide evidence for gastrointestinal infection of SARS-CoV-2, highlighting its potential fecal-oral transmission route.

---

Since the novel coronavirus (SARS-CoV-2) was identified in Wuhan, China, at the end of 2019, the virus has spread to 25 countries, infecting more than 68000 people and causing over 1600 deaths globally. Although a series of extraordinary social distancing measures have been implemented in China, the number of infections continues to rise. The viral infection causes a series of respiratory illness including severe respiratory syndrome, indicating the virus most likely infects respiratory epithelial cells and spreads mainly via respiratory tract from human to human. However, viral target cells and organs haven’t been fully determined, impeding our understanding of the pathogenesis of the viral infection and viral transmission routes. According to a recent case report, SARS-CoV-2 RNA was detected in a stool specimen^1^, indicating the possibility of the viral extrarespiratory infection and additional transmission routes to respiratory one. It has been proved that SARS-CoV-2 uses ACE2 as a viral receptor for entry process^2,3^. ACE2 mRNA is highly expressed in gastrointestinal system^4^, providing a prerequisite for SARS-CoV-2 infection. To further understand the clinical significance of SARS-CoV-2 RNA in feces, we examined the viral RNA in feces from 71 patients with SARS-CoV-2 during their hospitalization. Viral RNA and intracellular viral protein staining were also examined in gastrointestinal tissues from one of the patients.

## Methods

From February 1 to 14, 2020, clinical specimens including serum, nasopharyngeal and oropharyngeal swabs, urine, stool and tissues from 73 SARS-CoV-2-infected hospitalized patients were obtained in accordance with China Disease Control and Prevention (CDC) guidelines and tested for detection of SARS-CoV-2 RNA in 73 hospitalized SARS-CoV-2-infected patients using the China CDC-standardized quantitative polymerase chain reaction assay^5^. Clinical characteristics of the 73 patients were shown in Table 1. The esophageal, gastric, duodenal and rectal tissues were obtained from one of the patients using endoscopy. The patient’s clinical information was described in Supplementary Case Clinical Information and Supplementary table 1. Endoscopic overview images were shown in Supplementary Figure 1. Histological staining (H&E) as well as viral receptor ACE2 and viral nucleocapsid (NP) staining were performed as described in Supplementary methods. The images were obtained using a laser scanning confocal microscopy (LSM880, Carl Zeiss MicroImaging) and shown in Figure 1.

**Table 1.**
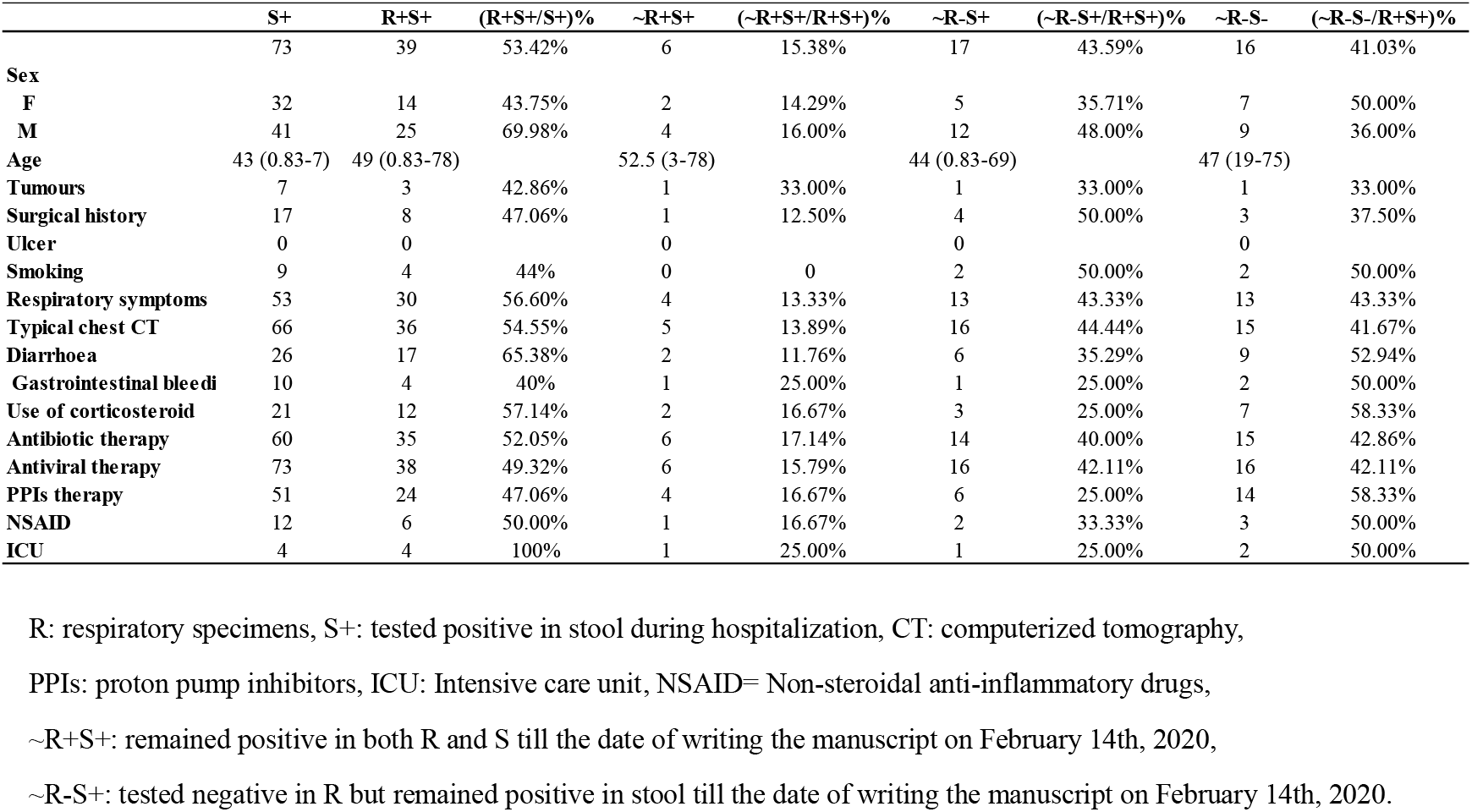
Clinical characteristics of patients with SARS-CoV-2 infection.

**Figure 1.**
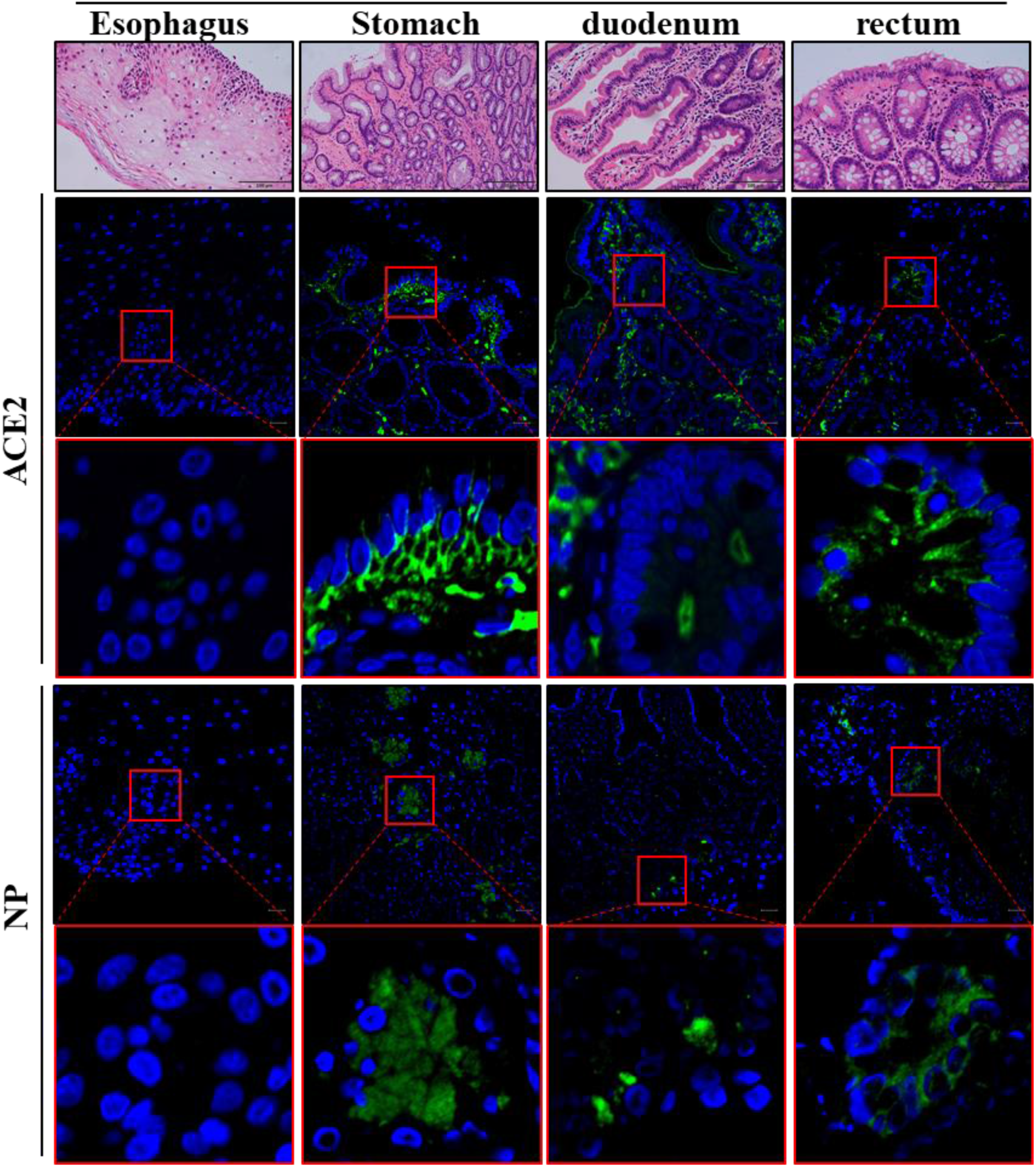
Images of Histological and Immunofluorescent Staining of Gastrointestinal Tissues. Shown are images of histological and immunofluorescent staining of esophagus, stomach, duodenum and rectum. The scale bar in the histological image represents 100 microns. The scale bar in the immunofluorescent image represents 20 microns.

## Results

From February 1 to 14, 2020, of all the 73 SARS-CoV-2-infected patients, 39 (53.42%) including 25 males and 14 females tested positive for SARS-CoV-2 RNA in stool (Table 1). The age of patients with positive SARS-CoV-2 RNA in stool ranges from 10 months to 78 years old (Table 1). Duration time of positive stool ranges from 1 to 12 days till the date of writing the manuscript on February 14, 2020 (Table 1). Furthermore, 17 (23.29%) patients remained positive in stool after showing negative in respiratory samples (Table 1).

Gastrointestinal endoscopy was performed on the patient described in Supplementary Material. Abnormality was not observed in the gastric, duodenum, colon and rectum except for mucosal lesions and bleeding in esophagus as described in Supplementary results (Supplementary Figure 1). All the gastrointestinal tissue samples obtained from esophageal, esophageal non-lesion, gastric, duodenum and rectum mucosa tested positive for SARS-CoV-2 RNA (Supplementary Table 1).

The mucous epithelium of esophagus, stomach, duodenum and rectum showed no significant damage with H&E staining (Figure 1). Infiltrate of occasional lymphocytes was observed in esophageal squamous epithelium (Figure 1). In lamina propria of stomach, duodenum and rectum, numerous infiltrating plasma cells and lymphocytes with interstitial edema were seen (Figure 1).

Importantly, viral host receptor ACE2 stained positive mainly in the cytoplasm of gastrointestinal epithelial cells (Figure 1). To note, we observed that ACE2 is rarely expressed in esophageal epithelium, but abundantly distributed in cilia of glandular epithelia (Figure 1). Staining of viral nucleocapsid protein (NP) was visualized in the cytoplasm of gastric, duodenal and rectum glandular epithelial cell, but not in esophageal epithelium (Figure 1).

## Discussion

In this manuscript, we provide evidence for gastrointestinal infection of SARS-CoV-2 and its possible fecal-oral transmission route. As SARS-CoV-2 continues to spread, it’s important to elucidate the viral transmission routes and take appropriate measures to control viral spread. Since viruses spread from infected to uninfected cells^6^, viral specific target cells or organs are determinants of viral transmission routes. Receptor-dependent viral entry is the first step of SARS-CoV-2 infection. Our immunofluorescent data showed that ACE2 protein, which has been proved to be the receptor of SARS-CoV-2, is abundantly expressed in the glandular cells of gastric, duodenal and rectal epithelia, allowing the entry of SARS-CoV-2 into the cells. ACE2 staining is rarely seen in esophageal mucosa probably because esophageal epithelium is mainly composed of squamous epithelial cells, while gastrointestinal epithelium below esophagus has abundant ACE2-expressed glandular epithelial cells.

Coronavirus genome encodes the spike protein, nucleocapsid protein, membrane protein and envelop protein to form a complete viral particle^7^. Beyond binding to viral genome to make up nucleocapsid, the nucleocapsid protein (NP) localizes to endoplasmic reticulum-Golgi region to facilitate viral assembly and budding^8^. Our results of viral RNA detection and intracellular staining of NP in gastric, duodenal and rectal epithelia demonstrate that SARS-CoV-2 infects these gastrointestinal glandular epithelial cells. Although viral RNA was also detected in esophageal mucous tissue, absence of NP staining in esophageal mucosa indicates low viral infection in esophageal mucosa probably due to lack of ACE2 protein expression. The data of viral protein staining are in line with the data of ACE2 staining, confirming the importance of ACE2 protein expression for SARS-CoV-2 infection.

After viral entry, virus-specific RNA and proteins are synthesized in the cytoplasm to assembly new virions^9^, which can be released to gastrointestinal tract. Recently, we and others have isolated infectious SARS-CoV-2 from stool (Manuscript under revision), confirming the release of the infectious virions to the gastrointestinal tract. Therefore, fecal-oral transmission could be an additional route for SARS-CoV-2 spread. Prevention of viral fecal-oral transmission should be taken into consideration to control the spread the virus.

The immune response to the viral infection of gastrointestinal tract needs to be further investigated. In this report, we observed infiltration of plasma cells and lymphocytes without obvious damage in gastrointestinal mucosa. The lesion and bleeding of the esophageal mucosa from a case of SARS-CoV-2 infection was probably stress-associated.

Our results highlight the clinical significance of testing viral RNA in feces by real-time reverse transcriptase polymerase chain reaction (rRT-PCR) since infectious virions released from gastrointestinal tract can be monitored by the test. According to the current CDC guidance for disposition of patients with SARS-CoV-2, the decision to discontinue Transmission-Based Precautions for hospitalized SARS-CoV-2 patients is based on negative results of rRT-PCR testing for SARS-CoV-2 from at least two sequential respiratory tract specimens collected ≥24 hours apart^10^. However, we observed in more than 20% of SARS-CoV-2 patients that the viral RNA remained positive in feces even after negative conversion of the viral RNA in respiratory tract, indicating that viral fecal-oral transmission can occur even after viral clearance in respiratory tract. Therefore, we strongly recommend that rRT-PCR testing for SARS-CoV-2 from feces should be performed routinely in SARS-CoV-2-infected patients, and Transmission-Based Precautions for hospitalized SARS-CoV-2-infected patients should continue if feces tests positive by rRT-PCR testing. In summary, our results provide evidence for gastrointestinal infection of SARS-CoV-2, which could result in fecal-oral transmission.

## Data Availability

The data used to support the findings of this study are available from the corresponding author upon request.

## Acknowledgements

Author Contribution: HS, FX design the study, analyzed the data and wrote the paper. FX, MT, XZ, CL, JH, and ZH contributed equally to this work.

